# Is the human face a biomarker of health? – a scoping review

**DOI:** 10.1101/2025.01.14.25320526

**Authors:** Weronika M. Obrochta, Paula Bartecka, Katarzyna Klaś, Magdalena Klimek, Urszula M. Marcinkowska

## Abstract

There is a widespread opinion that facial features could provide an important cue to individual’s health and are a biomarker of a human developmental stability. Taking evolutionary lens, they are interpreted to be a signal physical and cognitive health. However, research to date does not clearly support this assumption. This is the first review that explores the association between various aspects of health and facial features, namely symmetry, averageness or sexual dimorphism in adults. We searched electronic databases including Web of Science, MEDLINE PubMed, Scopus and Embase. We followed the Preferred Reporting Items for Systematic reviews and Meta-Analyses extension for Scoping Reviews (PRISMA-ScR) guidelines for reporting of our results. Of the 702 screened articles, 17 were eligible for inclusion. Studies presented a varied outcomes between facial features and cardiovascular health; immunocompetence; oxidative stress level; cortisol level; reproductive health, cognitive health and general physical health. This review presents mixed and inconclusive answer for the question whether facial features can serve as indicators of health. The results deepen our knowledge of the relationship between facial features and health outcomes and warrant caution when interpreting face as a biomarker of health. Protocol: Open Science Framework, https://osf.io/dv9pu/.

## Introduction

### Facial features in relation to health status

Facial appearance plays a crucial role in social interactions [1]. In line with theories of sexual selection, facial features can presumably act as indicators of an individual’s age, mating success, sexual behaviour and health [2]. Previous research has identified several facial features that may serve as indicators of health. These traits include e.g. symmetry, averageness, sexual dimorphism (masculinity and femininity), adiposity and skin colour. Among these, sexual dimorphism, averageness and symmetry have received extensive research attention [2]. Facial fluctuating asymmetry and averageness were suggested as morphological cues reflecting developmental stability and consequently being a proxy for individual’s biological condition [3]. Developmental stability is defined as the capability of an organism to sustain a consistent phenotype, despite potentially disruptive genetic and environmental factors encountered during prenatal and postnatal life. It is regarded as a vital component related to an individual’s survival and reproductive success [4]. Beneficial early-life environment is considered to be related with higher bilateral symmetry of facial traits, whereas pronounced asymmetries might indicate a disruption of developmental stability [5]. Sexually dimorphic features differ between an average, typical female and male phenotype of a given species. These secondary sexual traits are being shaped under the influence of sex-typical hormones, androgens and estrogens, and were suggested to be related to one’s health and reproductive potential [6–9].

### Facial fluctuating asymmetry

Fluctuating asymmetry (FA) refers to random, small deviations from perfect bilateral symmetry, typically calculated across multiple traits. It is also regarded as an indicator of an organism’s capacity to withstand or mitigate environmental disturbances during the prenatal development [10, 11]. For individuals manifesting minimal deviations from ideal symmetry, low FA indicates potential developmental stability and high biological quality [12]. However, in the up-to-date studies results for FA as a marker of health are mixed and inconclusive [13, 14].

### Facial averageness

Facial averageness, next to symmetry, is thought to reflect an individual’s ability to withstand the negative impacts of genetic and environmental stressors during development [2, 15, 16]. Facial averageness is also often associated with attractiveness [17–19]. What is more, it was found that individuals with average traits have higher fitness (i.e. higher number of offspring). Therefore, facial averageness could also signal reproductive health [20]. On the other hand, facial distinctiveness can be considered as the opposite of averageness, and its relationship with actual health has also been examined. A negative correlation was observed between facial averageness and semen quality; however, no association was found between averageness and immune function [2]. An inverse relationship between distinctiveness and other aspects of measured health were also observed [13]. Nevertheless, similarly to asymmetry, results of the published studies are incongruent [9, 21, 22].

### Facial sexually dimorphism

More masculine or more feminine facial features were previously suggested as indicators of sex hormone exposure during development [23]. More masculine facial features are manifested in strong jaw, brow ridge and higher facial weight-to-height ratio [24]. Facial masculinity may convey information about health through its association with testosterone [25]. Testosterone, while enhancing overall masculinity or muscle mass, can also compromise health by suppressing immune functions [21] and elevating oxidative stress levels [26]. Testosterone also plays a vital role in spermatogenesis [27], suggesting that masculinity may also be related with semen quality [2]. More feminine facial features are defined as fuller lips, bigger and rounder eyes, and narrow chin. Facial femininity has been suggested to be linked with estrogens [28], that also play a role in enhancing immune function [29] and reproductive health, given that higher levels of estradiol are needed for a successful ovulation [30, 31]. Nevertheless, the impact of sex hormones on women’s health remains a topic of debate and is possibly not as strong as in men [21].

### Cognitive and computational approaches to facial features

Studies included in the current review were designed employing either cognitive or computational approach. The cognitive approach to assessments of facial features involves participants evaluating facial appearance of presented facial stimuli [32]. Evaluators are randomly assigned to grade facial photographs on one of the chosen traits e.g. perceived health or attractiveness, via Alternative Forced Choice or on a Likert Scale. For each trait, the average score for designated face is calculated by averaging all evaluators’ ratings[2], sometimes accounting for the within and between rater agreements [33, 34].

The computational approach is based on measuring facial features from photographs. This method involves applying landmarks to the facial photographs and establishing their coordinates on a 2- or 3-dimensional grid. It is worth mentioning the importance of standardisation of photographs, i.e. employing Frankfurt horizontal plane (participant’s head position, with the camera set at his or her eye level) [35], constant lightening and distance between camera and participants. To standardize the location, orientation and scale of landmarks and semi-landmarks configurations are employed, i.e. via superimposed generalised Procrustes analysis. The digitizing process is frequently carried out in the program tpsDig2 or Geomorph package [22, 35], but previously also other methods were employed [36]. The landmarking can be done manually or automatically (with slight differences between two techniques) [37].

### The aim of the scoping review

So far, no review has been conducted that would comprehensively describe the up-to-date literature on the association between the facial features and health status. Our review fills this gap. It is of great importance, as frequently the connection between facial features and underlining “biological quality” is assumed and treated as a given. This stand is reinforced by the evolutionary rationale for bases of what humans find attractive (and why attractiveness to e.g. symmetry or sexual dimorphism would be adaptive) [38]. This scoping review offers a summary of findings on the relationship between health and facial features (symmetry, averageness and sexual dimorphism), while simultaneously stratifying the results by types of health measurements, and accounting for the employed approach in facial features evaluation (cognitive or computational).

## Materials and methods

### Protocol, Registration, and Reporting Methods

We conducted this scoping review according to JBI (formerly Joanna Briggs Institute) methodology for scoping reviews [39] guided by a protocol registered in Open Science Framework (https://osf.io/dv9pu/). We reported this review in accordance with the Preferred Reporting Items for Systematic Reviews and Meta-Analyses extension for scoping reviews (PRISMA-ScR, see S1 Table) [40].

### Search strategy

We implemented the three-step search strategy endorsed by JBI for conducting scoping reviews [39]. To build our search strategy, we conducted a series of pilot searches of the relevant databases to identify articles focused on the topic and appropriate keywords. An initial pilot search of Web of Science, MEDLINE PubMed, Cochrane Database and Embase was developed by the first author (WO). The analysis of terminology within the articles allowed to develop a full search strategy. Records obtained from the pilot search were reviewed to confirm the appropriateness of the used keywords to identify articles related to the scoping review’s research questions. Based on that we developed the following search strategy: *“health” AND (facial AND ((symmetry OR averageness OR dimorphism) NOT (attractiveness OR palsy))) NOT children*.

The databases searched for the final analysis were MEDLINE PubMed, Web of Science, Scopus and Embase. The final search was conducted on March 1, 2024. Two independent judges (WO and PB) screened the reference lists of all included articles for further studies.

### Eligibility criteria

***Concept and Context.*** The concept of interest focused on studies evaluating the association between health status and chosen facial features. We included studies if 1) they examined facial symmetry, averageness or sexual dimorphism in relation to health, 2) they assessed health through analysis of biological material (i.e. blood samples, saliva samples), medical records or questionnaires (self-report, ratings). We included studies with computational and cognitive approaches to assessing facial features. We excluded articles reporting on participants’ health measured retrospectively e.g. a survey about health state in childhood. ***Participants***. We included studies involving individuals over 18 years old. We excluded studies that report results of individuals with a history of craniofacial surgeries, Bell’s palsy, lip cleft, or current or past extensive orthodontic treatment. ***Types of Sources***. The scoping review considered observational and quantitative studies that describe relation between facial features and health. Qualitative studies, case reports, books, letters and correspondence were excluded. ***Publication date***. There were no restrictions on the publication dates of articles other than the search timing. ***Language***. Only studies published in English were included in the review.

### Study Selection

Following the search, all identified papers were collected and uploaded into EndNote 21.2.0.17387 (2023). Duplicates were removed. Titles and abstracts were screened by two independent reviewers (WO and PB) for assessment against the eligibility criteria. The full texts of selected articles were assessed in detail against the eligibility criteria by two independent reviewers (WO and PB), see Fig 1 for more details. Data was screening by two independent reviewers (WO and PB) using a data screening tool – Rayyan [41]. The reviewers resolved any disagreements that arose between through discussion or with additional participation. of experienced reviewers (UMM and MK).

**Fig 1.**
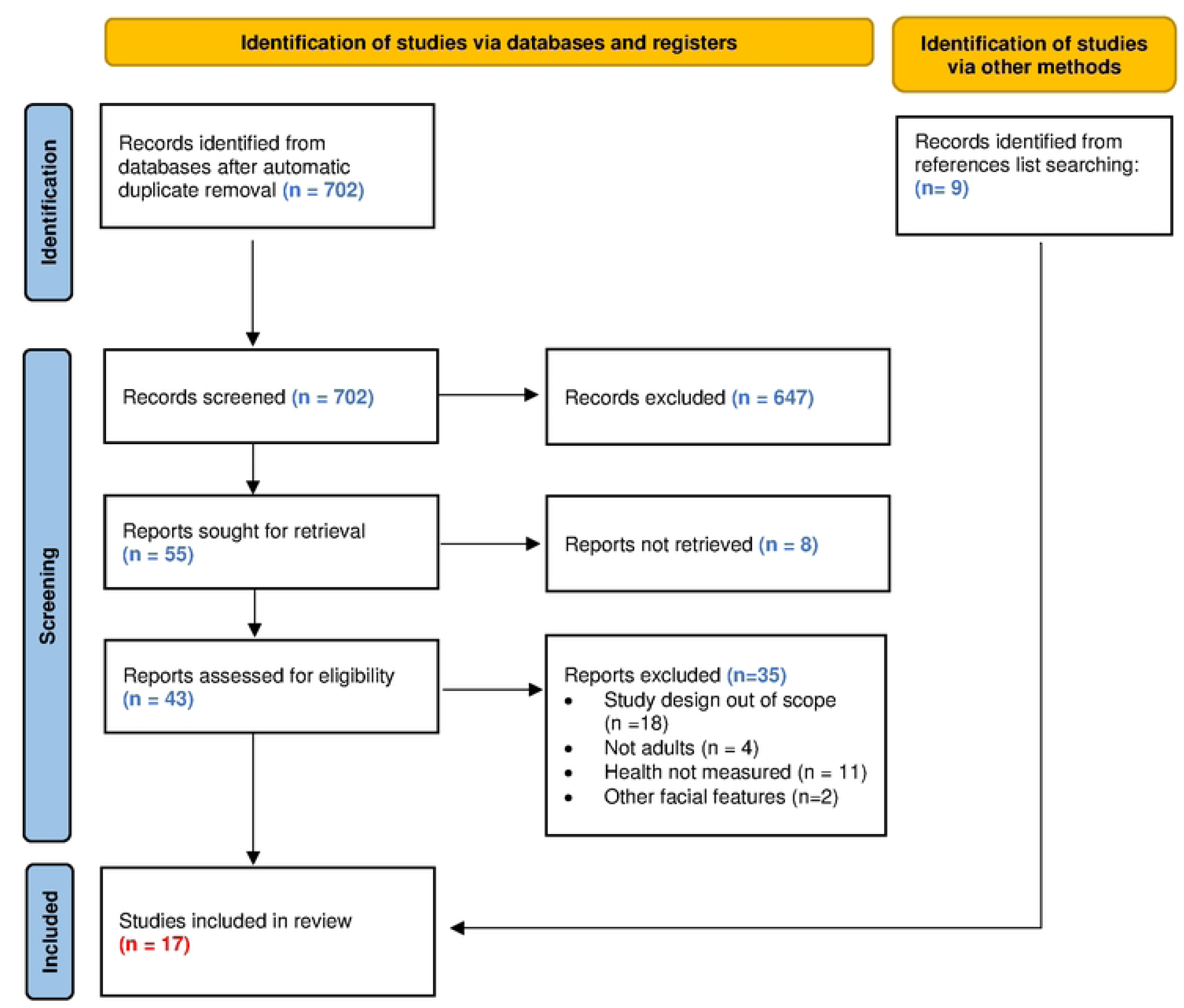
PRISMA 2020 flow diagram. * consider, if feasible to do so, reporting the number of records identified from eachdatabase or register searched (rather than the total number across all databases/registers). ** If automation tools were used, indicate how many records were excluded by a human and how many were excluded by automation tools.

### Data Extraction

We extracted details about the participants, concept, context, study methods and key findings relevant to the review questions. Two reviewers (WO and PB) independently gathered the data from each study and extracted information on relation between facial features and health status and employed methodology.

### Quality assessment

Quality assessment was not conducted, as per the JBI guidance on scoping reviews [39].

### Data synthesis and analysis

Public health studies are often complex as measuring human health holistically is virtually impossible. They investigate diverse populations and are characterised by heterogeneous endpoints or varied methodological approaches. As a result of the complexity, significant heterogeneity in analysed research was expected.

We used tables for synthesis, and descriptive summaries were utilised for reporting on the article characteristics. Two reviewers (WO and PB) independently and then collaboratively (with UMM and MK) summarised the results’ tables. We compared similarities based on the facial features studied and types of health measurements. Due to the multiple approaches to health measurements, for the sake of scientific clarity we developed categorisation guide to segregate results as follows: 1) cardiovascular health; 2) immunocompetence; 3) oxidative stress level; 4) cortisol level; 5) reproductive health 6) cognitive health; 7) general physical health.

## Results

### Literature search

A total of 702 titles and abstracts were reviewed, and 43 articles were identified for a full-text review. There were 35 qualitative studies that were excluded (see Fig 1 for exact reasons for exclusion). We additionally included 9 records identified manually from references lists search, resulting in 17 articles included in the final analysis (Fig 1, Table 1). The 17 articles considered the results of 24 analyses.

**Table 1.**
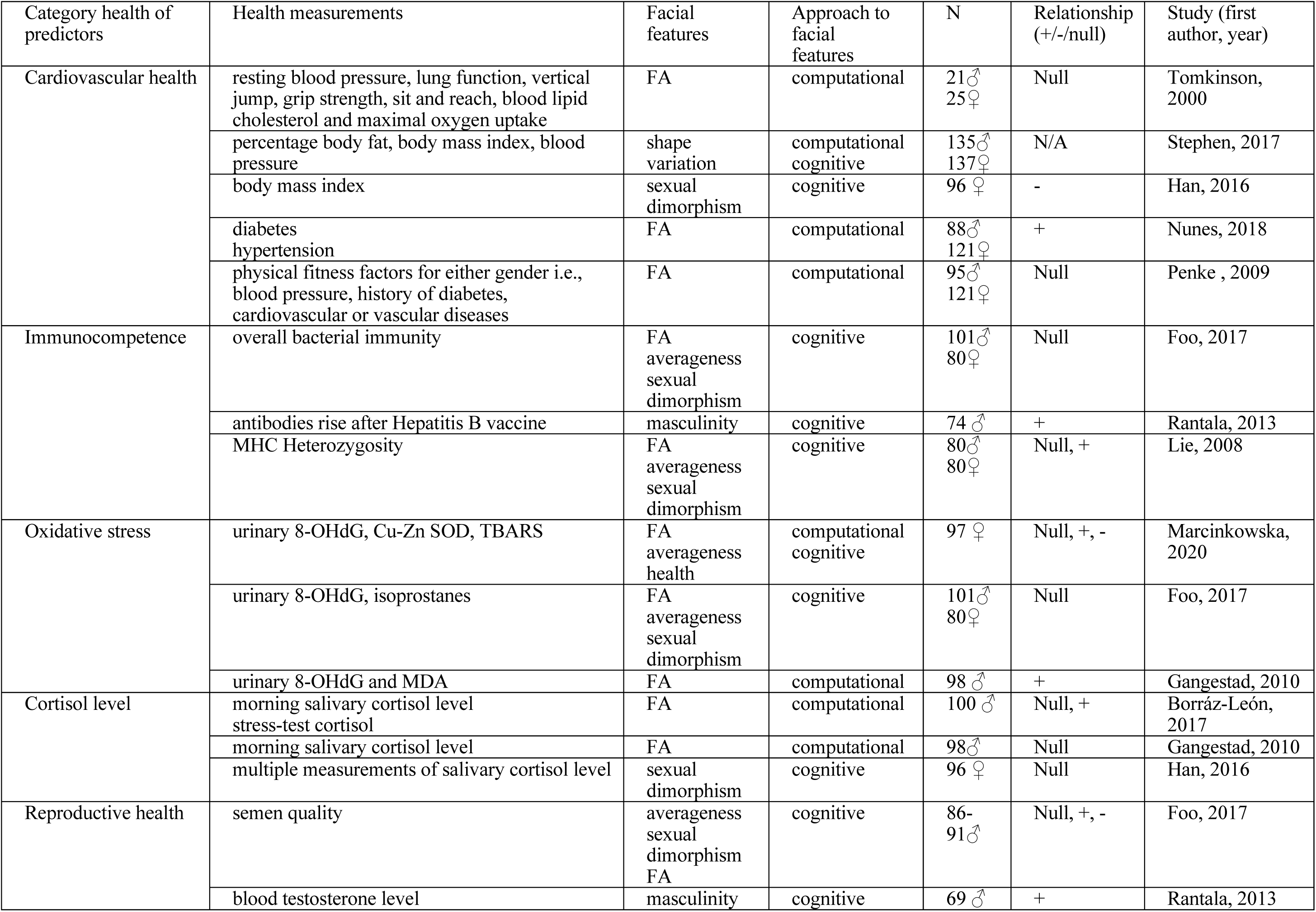
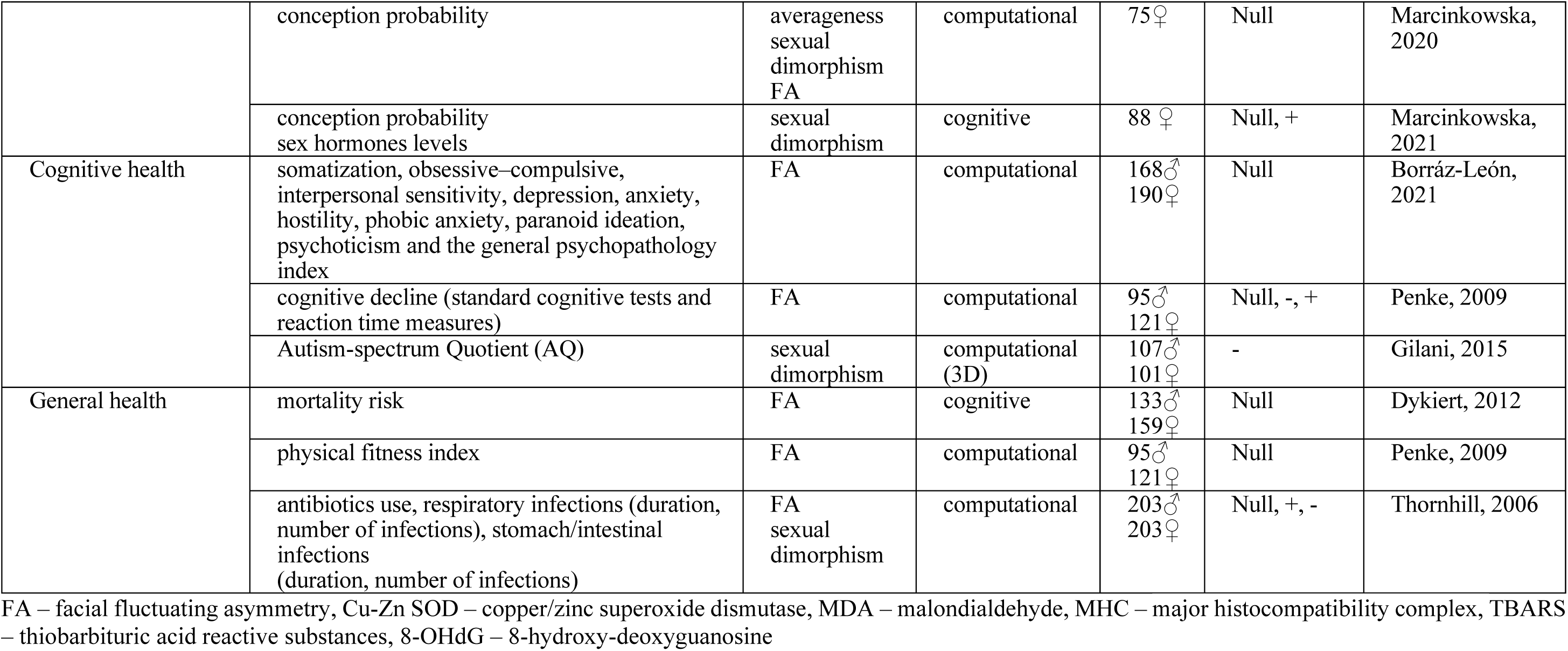
Studies reporting on cardiovascular health and facial features.

### Articles characteristic

The earliest publication identified was published in 2000 by *Tomkinson et al.* [5] The latest publications included in this review was *Borráz-León et al.* [42] and *Marcinkowska et al.* [43] both published in 2021. Around half of the studies (9/17; 53%) were published since 2015, with vast majority published since 2010 (15/17; 88%, Table 1). Most of the included articles evaluate facial fluctuating asymmetry or symmetry (11/17; 65%), and facial femininity or masculinity (11/17; 65%), and only few evaluated facial averageness (3/17; 18%).

### Facial features and various aspects of health

#### 1. Cardiovascular health

*Stephen et al.* [17] measured facial shape variation (expressed as a combination of facial symmetry, averageness and sexual dimorphism) and its’ relation to cardiometabolic health with both computational and cognitive approaches. They employed geometric morphometric method [44] of facial landmark data to forecast the presence of risk factors for cardiovascular disease – Body Mass Index (BMI, 32% of variance explained by facial shape), percentage of body fat (21% explained), and blood pressure (21% explained). Furthermore, the BMI and blood pressure values, although not a percentage of body fat exhibited a significant correlation with health judged by random judges (rated facial health). In a study by *Han et al.* [45], BMI was found to be negatively related to facial femininity (r=–0.32, p=0.002). *Nunes et al.* [46] used geometric morphometrics approach to identify facial shape traits associated with the presence of diabetes, hypertension or both conditions (D2 Mahalanobis distances ranged between 1.78-6.10; all p-vales <0.02). The largest facial morphological disparity was observed between individuals without these conditions and those with diabetes. Additionally, individuals with hypertension tended to exhibit higher levels of facial asymmetry (p>0.05, effect sizes not provided). Incongruently, *Tomkinson et al.* [5] reported no significant relationship between health-related physiological parameters (resting blood pressure, lung function, vertical jump, grip strength, sit and reach, blood lipid cholesterol and maximal oxygen uptake using cycle ergometry, correlation coefficients between −0.41 and 0.32, ps>0.05) and facial asymmetry. Similarly, *Penke et al*. [47] did not report significant relationships between horizontal fluctuating asymmetry (HFA) and comprehensive fluctuating asymmetry (CFA) indices and the physical fitness factors for either gender i.e. blood pressure, history of diabetes, cardiovascular or vascular diseases (rs<|0.11|, ps>0.30).

#### 2. Immunocompetence

Cognitive approach results in *Foo et al.* [2] indicated weak associations between facial features and various aspects of immune function. Multiple regression models showed that bacterial immunity was not related to facial appearance predictors (p-values ranged between 0.20–0.66) in either men or women. Going further, *Rantala et al.* [48] reported an association between hepatitis B antibody response and facial features. They found that following a hepatitis B vaccination protocol, there was a positive association between men’s immune reaction and facial masculinity (r=0.47, p<0.001) rated by female participants. On the other hand, *Lie et al.* [34] did not find a significant relationship between facial symmetry or masculinity judged by opposite-sex raters and an indirect measure of innate immunity (diversity at the major histocompatibility complex, MHC) in men. However, the study revealed that facial averageness was associated positively with overall MHC heterozygosity in men (R2=0.177; p<0.001).

#### 3. Oxidative stress level

Oxidative stress (OS), among other factors, contributes to the development of diabetes, obesity, and diabetes-related microvascular diseases, atherosclerosis, and cardiovascular diseases [49, 50] and is frequently used as a biomarker of the condition of an organism, its health and pace of ageing. *Marcinkowska et al.* [22] showed mixed results, including positive, negative and null relationship between OS and facial features in a sample of postmenopausal women. In this study a multivariate regression analysis was used to investigate the relationship between OS levels (measured by DNA damage and 3 levels of biomarkers, namely 8-hydroxy-20-deoxyguanosine (8-OHdG), copper/zinc superoxide dismutase (Cu-Zn SOD), thiobarbituric acid reactive substances (TBARS) levels) and facial morphology features calculated using PsychoMorph Program [51]. Analyses were based on computational and cognitive approaches. Interestingly, the first analysis with a cognitive approach showed that faces of women with high OS were chosen as less symmetrical (F=-4.370, p=0.037), but healthier (F=84.39, p<0.001), than faces of women with low OS. The second analysis, where the Geometric Morphometric Modelling was used to calculate the facial features, did not confirm any statistically significant relationship between OS biomarkers and facial morphology (all p-values >0.449). In the third study, based on a computational approach, negative correlations between both facial symmetry (b=-0.096, 95% CI [−0.18, −0.01], R2=0.042, p=0.025) and averageness (b=-0.140, 95% CI [-0.25, −0.03], R2=0.050, p=0.016) and 8-OHdG were observed. Moreover, Cu–Zn SOD, was negatively correlated with averageness (b=-0.054, 95%CI [−0.10, −0.01], R2=0.038, p=0.032) but not symmetry. No statistically significant associations with either averageness or symmetry and TBARS were detected [22].

*Gangestad et al.* [52] also explored 8-OHdG as a marker of OS levels and additionally malondialdehyde (MDA), which is a marker of lipid oxidative damage. In their computational approach, FA significantly and positively predicted levels of urinary OS biomarkers (r=0.26, p=0.021 for aggregated OS biomarkers, r=0.24, p<0.05 for 8-OHdG, and r=0.16, no p value reported for MDA). Moreover, the study found no evidence that either cortisol or testosterone mediate association between FA and OS levels. On the contrary, *Foo et al.* [2], showed no significant relationship between facial features and OS measures (urinary 8-OHdG and isoprostanes) in women and men (0.16<ps<0.91).

#### 4. Cortisol level

Cortisol is a steroid hormone that performs various roles in the human body, including managing the stress response, regulating metabolism, the immune function and inflammatory response [53, 54], and is frequently used as a biomarker of chronic stress exposure and health deterioration resulting from it. Analyses based on facial measurements in *Borráz-León et al.* [55] showed no significant associations between facial FA and morning salivary cortisol levels (p>0.05). However, after a stress test, symmetrical men (with lower FA) showed an increase in cortisol levels, whereas asymmetrical men (with higher FA) exhibited a decrease in cortisol levels (b=0.788, p<0.001). Additionally, *Gangestad et al.* [52] showed that there was no relationship between cortisol level and FA (p>0.05). Likewise, *Han et al.* 2016 [45] did not observe significant correlations between cortisol levels (averaged over 5 test sessions) and ratings of facial femininity in women (r=-0.08, p=0.43).

#### 5. Reproductive health

*Foo et al*. [2] measured relation between facial features and semen quality in men. They indicated that the linearity of sperm movement was positively predicted by facial masculinity (r=0.29, p=0.01). Furthermore, sperm concentration and percentage of motile sperm was negatively predicted by facial averageness (r=-0.21, p=0.04) and positively by facial masculinity rated by opposite-sex judges (r=0.23, p=0.03). *Rantala et al.* [48] showed that facial masculinity was significantly correlated with testosterone levels (r= 0.38, p=0.001) in men. On the other hand, a study of *Marcinkowska et al.* [56] found no evidence for a change in facial asymmetry, averageness or sexual dimorphism between three different points in the menstrual cycle that vary in conception probability and levels of female typical sex hormones, namely estradiol and progesterone (Fs≤0.78, partial η2s≤0.01, ps≥0.542). In another study, *Marcinkowska et al.* [43] using complex methodological cognitive approach (comparison of results from 3-Alternative Forced Choice and Likert Scale judgements, inclusion of varying visual stimuli and based on a cross-cultural sample of raters), found limited evidence for the relation between femininity, and conception probability and sex hormones. Even more importantly, the few statistically significant effects differed depending on the methods employed - forced choice vs. Likert Scale. Strikingly, associations between two methodological approaches were statistically significant (p=0.046) in 1 out of 5 analyses. This result provides methodological evidence for complexity of the relationship between facial judgement and facial features.

#### 6. Cognitive health

*Borráz-León et al.* [42] showed no correlations (in a mixed group of both men and women) between facial symmetry and minor mental ailments i.e. Somatization, Obsessive–Compulsive, Interpersonal Sensitivity, Depression, Anxiety, Hostility, Phobic Anxiety, Paranoid Ideation, Psychoticism, and the General Psychopathology Index measured via Symptom Checklist-90-Revised (SCL-90-R), rs<0.10, ps>0.05). *Penke et al.* [47] showed that facial asymmetry indices (namely horizontal fluctuating asymmetry, HFA; and comprehensive fluctuating asymmetry index, CFA) at age 83 were unrelated to measured intelligence assessed at age 11, 79 or 83 (all p-values >0.05). Facial asymmetry at age 83 was also not related to change in cognitive abilities from age 11 to age 79 years (p>0.05). However, cognitive decline between age 79 and 83 in men was significantly, negatively related to all symmetry indices (rs=−0.24 to −0.35, 0.001<ps<0.05); men with lower FA at age 83 had experienced less cognitive decline in the preceding 4 years, and showed lower reaction times (rs=.05 –.30) This effect was not replicated in women. *Gilani et al.* [57] showed that men and women with high levels of autistic-like traits present less prominent facial sexual dimorphism than individuals with low levels of autistic-like traits (for 4 out of 6 facial measurements depending on the sex, for men <0.001<ps<0.05; for women <0.001<ps<0.003). One of the measurements (nasal bridge length) showed an opposite pattern in women – shorter average nasal bridge length (more feminine) was observed in a group of higher autism scores.

#### 7. General health

*Dykiert et al.* [58] indicated that rated facial symmetry was not associated with the risk of mortality in a group of participants with an average age of 83 years, who were monitored over a period of 7 years (p=0.55 for the whole sample; p=0.32 for men; p=0.58 for women). Also, *Penke et al.* [47] did not report significant relationships between any of the two measured symmetry indices (HFA, CFA) and the physical fitness factors for either sex (rs<|0.11|, ps>0.30). *Thornhill & Gangestad* [59] demonstrated that facial masculinity interacted with sex and predicted the number of reported occurrences of antibiotic use (measure of effective immunocompetence) and respiratory infections (for number of infections beta=-0.19, p=0.001, for days of infection beta=-0.17, p=0.002, effect stronger for males than for females). No such association was observed for stomach/intestinal infections (beta=0.04, p>0.05). Moreover, FA was not related to the total number of infections (p=0.139), and only marginally predicted the total days infected (p=0.07). On the other hand, FA was positively associated with days (beta=0.14, p=0.011) and number (beta=0.11, p=0.03) of respiratory infections. It is worth mentioning that the association between FA and number of respiratory infections was only marginally significant when controlled for potential confounders. No effect was observed when FA and intestinal ailments were analysed (p>0.05). Facial symmetry was marginally related to the number of times that antibiotics were used (p=0.057) for both sexes combined.

## Discussion

This scoping review provides a comprehensive evaluation of literature focused on the relationship between most frequently analysed facial features (asymmetry, averageness and sexual dimorphism) and various measures of health, including cardiovascular, reproductive and overall health, oxidative stress level and immune function. Across 17 articles included in the review, 24 analyses on facial features and health were conducted (frequently one article reported results on multiple aspects of health). Of the five studies including the cardiovascular health, only one showed a positive relationship with facial features. Positive correlation with immunocompetence was also depicted in one study out of three and partially in a second one. When it comes to oxidative stress and reproductive health and facial features the analyses present all possible results (positive, negative and null). None of the studies found a relationship between FA, femininity and salivary cortisol level, aside from one study showing a relationship between C reaction to stress and FA. Of the three studies, two showed negative relationship with cognitive health (Table 1). Three studies tested relation between general health measures and facial features, and all reported null results (either entirely, or partly). Although there seems to be an assumption that facial features are a signal of healthiness, and face as such seemed to be interpreted as a biomarker of health and biological quality, the current body of evidence does not provide a strong support for this claim.

Other than the actual lack of relation between facial features and health in some studies, there can be multiple reasons for discrepancy in the results. The articles included in the review included adult populations from Europe, Asia, Africa and the Middle East. Participants in the study ranged in age from 18 to 83. Fourteen studies involved male as well as female participants, 6 focused only on male participants and 5 only on females. The pronounced variation on the sampling level could have led to varying results and effect sizes of the reported significant effects. Additionally, the differences between publications could stem from different methods of measuring facial features and also due to a great array of employed measures of health. Computational approaches (measuring the facial features) differed in protocols and number of landmarks. The cognitive approach (judgements of the facial features) in most, but not all publications relied on opposite sex raters. Also, most studies did not report between and within rater-agreements.

### The significance of attractiveness and facial adiposity

When considering face as a biomarker of health, two more aspects seem to be closely related, facial attractiveness and adiposity. As facial attractiveness refers to other people’s perception, it can only be assessed by cognitive approach. If face was a biomarker of health, then facial appearance could serve as an honest signal manifesting mate quality to potential mates. Taking the evolutionary lens, individuals who would find “healthy faces” attractive would then be able to obtain better genes or/and improved fitness for their children. Published studies present mixed results for the relationship between the three facial characteristics of interest (sexual dimorphism, averageness, and symmetry) and attractiveness. Although theories of sexual signaling predict that attractive appearance would be positively related to actual health [2, 60, 61], not all studies found such connection.

*Foo et al.* [2] found that neither symmetry nor averageness significantly predicted perceived attractiveness in females. On the other hand, *Rantala et al.*[48] showed that antibody response was significantly correlated with facial attractiveness (r=0.43, p<0.001). Similarly, *Lie et al.* [34] showed that MHC heterozygosity positively predicted male attractiveness [34]. *Gangestad et al.* [52] found a modest negative correlation between male physical attractiveness and OS biomarkers levels, but *Foo et al.* failed to find such relationship [2]. Interestingly, *Marcinkowska et al.* reported that faces of women with high OS were perceived as more attractive [22]. There are also mixed results on the relationship between attractiveness and plasma cortisol level [54, 62]. In the overall health domain, *Thornhill & Gangestad* presented no associations between facial attractiveness and total infections and antibiotic use [59]. No correlations were found between minor ailments in mental health outcomes and other-perceived attractiveness, however, self-perceived attractiveness was a significant predictor of health among both men and women[42]. Attractiveness also did not significantly predict mortality [58]. These strongly mixed findings suggest that even if facial symmetry and averageness would indicate health, the perception of attractiveness based on these cues may not be as universally consistent as previously expected [2, 56]. Importantly, if the components of attractiveness were symmetry, averageness and sexual dimorphism, then evidence for the interpretation of attractiveness as an adaptive signal of health is not as strong as assumed.

Another theoretical concept that is closely related to facial appearance and health is adiposity. *Foo et al.* showed that male perceived health was negatively predicted by adiposity [2]. A study by *Coetzee et al.* indicated that facial adiposity can act as a cue to health in young adult participants [63]. Research showed also a negative correlation between adiposity and immune response in men [48]. Interestingly, women’s adiposity did not correlate with immune responsiveness [62]. The review of *de Jager et al.* 2018 presented 3 studies which investigated the relationship between facial adiposity and mental health. Only one of these three articles found that rated facial adiposity was negatively correlated with a psychological condition factor in women [64, 65]. Due to the possible relationship between adiposity and health, and adiposity and facial appearance, future studies should include a measurement of it as a confounding factor.

### Strengths and limitations

This scoping review can serve as a valuable resource that charts the existing evidence for linkage between health and facial shape, offering the characteristics of the included studies, highlighting gaps and areas requiring further research. The strength of this scoping review is the standardized literature screening process and the evaluation of the extracted data by the second and third reviewers, which increases the credibility and validity of the review. The scientific transparency and open science approach (https://osf.io/dv9pu/) provides a foundation for subsequent systematic reviews or primary research to investigate the evidence on the relationship between health and facial features, employing various measurement approaches across different populations and settings.

Nevertheless, there are several limitations to our scoping review that should be noted. Firstly, the search strategy’s scope, either involving restrictive inclusion/exclusion criteria and the selected databases may not encompass all relevant literature on the research topic, objectives, and questions. Secondly, language restrictions by focusing solely on articles published in English could omit evidence published in other languages. There is a risk that relevant articles in other languages were excluded. In addition, we have not included “grey literature” i.e. evidence published in forms other than scientific articles. Finally, due to the high heterogeneity of the studies, it is currently not possible to carry out a meta-analysis that could provide a numeric answer to the validity of face as biomarker approach. Possibly, with accumulation of new published studies in future a meta-analytic approach will be possible.

### Conclusion

This review verifies a common assumption that the face is a biomarker of health [1]. However, most of the studies to date on this topic have not confirmed this assumption. Out of 24 analyses that were retrieved from the existing 17 articles, as many as 17 showed null results, either entirely or partly. Some of the studies also found results opposite than expected (worse health was related to better facial judgements or measurements, i.e. higher OS was related to greater perceived facial health [56]. Other than methodological and sampling differences that could have led to the pronounced results’ incongruency, it is also plausible that the previously widely assumed interpretation of face as a biomarker of good health and beneficial developmental conditions actually lacks a scientific, evidence-based grounding.

More studies are needed that comprehensively test the relationship between facial features and physical and cognitive health, especially in the areas were results are most incongruent, i.e. immune health, oxidative stress and reproductive health.

Additionally, with more studies employing standardised approaches based on publicly available protocols, it might be possible to either rule out previous hypotheses or to add more knowledge to support its scientific grounding, and as an outcome understand the complexity that led to the current lack of agreement on whether facial features can serve as a biomarker of health. Currently, we cannot conclude that there is sufficient support for the hypothesis that the human face contains valid cues to health, and that facial appearance therefore provides a reliable cue for identifying healthy and unhealthy individuals.

## Acknowledgments

Not applicable.

## Declarations

### Author contributions

Conceptualization: WO, UMM, MK Methodology: WO, KK Investigation: WO, PB, UMM Visualization: WO, KK, MK Supervision: KK, MK Writing—original draft: WO, PB, UMM Writing—review & editing: WO, KK, MK, PB, UMM

### Data availability

Data and material are available from the authors upon request to the corresponding author.

### Patient and public involvement

Patients and/or the public were not involved in the design, or conduct, or reporting, or dissemination plans of this research.

### Patient consent for publication

Not applicable.

### Supporting information

S1 Table. Preferred Reporting Items for Systematic reviews and Meta-Analyses extension for Scoping Reviews (PRISMA-ScR) Checklist

